# Self-Stigma and Depression Among Community-Dwelling Adults With Physical Disabilities in China: The Chain Mediating Role of Social Participation and Self-Esteem

**DOI:** 10.1101/2025.11.23.25340835

**Authors:** Qianqian Hu, Jingjing Gong, Aixiang Li, Rui Wang, Hui Chen

## Abstract

**Background:** With the intensification of the aging population, the number of people with physical disabilities in China is gradually increasing. Physical disabilities have led to the formation of unique psychological characteristics among this group, seriously affecting their thoughts, behaviors, and quality of life. Therefore, this study explores the action paths and predictive values of self-stigma, social participation, self-esteem, and their influence on depression among community-dwelling people with physical disabilities in China based on the modified labeling theory.

**Methods:** From April to September 2023, a cross-sectional study was conducted on 280 people with physical disabilities recruited from 8 counties and cities in Yanbian Prefecture, China. Various dimensional scales were used to measure depression, self-stigma, social participation, and self-esteem. SPSS 26.0 was employed for descriptive statistical analysis, and Amos 26.0 was used to construct a structural equation model and analyze the mediating effect.

**Results:** The average score of depression was (43.08±9.28). Pearson analysis showed that self-stigma, social participation were significantly positively correlated with depression, while self-esteem was significantly negatively correlated with depression (all *p*<0.05). Path analysis results indicated that self - stigma could directly affect depression (*β*=0.297,*P*<0.001), and also indirectly affect depression through social participation (*β*=0.265, *P*<0.01), self-esteem (*β*=0.106, *P*<0.01), and both social participation and self-esteem (*β*=0.076, *P*<0.01), with effect sizes of 39.92%, 35.62%, 14.25%, and 10.22% respectively.

**Conclusion:** Social participation and self-esteem play a chain-mediating role between self-stigma and depression among community-dwelling people with physical disabilities. This provides a reference for formulating social intervention plans to improve the mental health of people with physical disabilities.

## 1. Introduction

Depression is a state characterized by low mood and aversion to social activities, which affects an individual’s thoughts, behaviors, and quality of life. It manifests as sleep disorders, decreased appetite, and may even lead to extreme ideas and behaviors such as self-harm and suicide^[1,2]^. According to surveys, 40% of the 280,000 suicide cases that occur in China each year involve individuals suffering from depression^[3]^. People with disabilities are at a disadvantage in aspects of social life. They often face unfair categorization from the outside world and are regarded as a “mentally incompetent” and “disabled” group that is useless to society^[4]^. In Link’s modified labeling theory, it is posited that once an individual is classified as a member of a particular special group and anticipates being devalued or discriminated against, negative social perceptions increase the individual’s sensitivity to self-negative evaluation or rejection, triggering self-protection mechanisms and prompting some defensive coping measures^[5,6]^. For example, they may avoid or reject potential uncomfortable or threatening social interactions. This results in reduced income, a smaller social network, and an increased likelihood of recurrence or exacerbation of mental illnesses^[7]^.

Influenced by stigma, the disabled population internalizes these negative labels imposed by the outside world as part of their self-evaluation, and forms self-stigma through identification and application. These negative stereotypes and labels subject them to unfair treatment such as prejudice and discrimination in aspects like daily life, employment, education, and access to medical services^[8]^. These experiences lead them to change their self-perception, intensify their sense of frustration and shame in interpersonal interactions, and result in low self-esteem, reduced willingness to participate in society, and feelings of isolation and depression^[9]^.

Self-esteem is one of the core elements in personality development and represents an individual’s subjective evaluation of the self. Previous studies have indicated that self-esteem plays a role in buffering depression, and its level affects an individual’s mental health, interpersonal communication, personality integrity and stability, as well as subjective initiative^[9]^. Social participation is an important indicator reflecting public health. Individuals with low levels of social participation tend to have lower quality of life and poorer physical and mental health^[10,11]^.

Currently, social participation has not been incorporated into relevant research in China. The specific influence paths and effects among self-stigma, self-esteem, social participation, and depression remain unclear. Therefore, this study aims to explore the current level of depression among community residents with physical disabilities based on the modified labeling theory, clarify the path relationships among self-stigma, self-esteem, social participation, and depression, and provide a basis for community workers and primary healthcare institutions to carry out targeted psychological nursing interventions for community residents with disabilities, so as to improve their mental health levels.

### 1.1 Research Hypotheses

H1: There is a positive correlation between self-stigma and depression.

H2: There is a negative correlation between social participation, self-esteem and depression.

H3: There is a negative correlation between self-stigma and social participation, self-esteem.

H4: Social participation plays a mediating role in the relationship between self-stigma and depression.

H5: Self-esteem plays a mediating role in the relationship between self-stigma and depression.

H6: Social participation and self-esteem play a chain-mediating role in the relationship between self-stigma and depression.

## 2 Methods

### 2.1 Participants

A cross-sectional study was conducted using the convenience sampling method in eight counties and cities of Yanbian Prefecture, Jilin Province, China. From April to September 2023, a total of 280 eligible community residents with physical disabilities were recruited to participate in this study.

Inclusion criteria:(i) Aged 18 years or older; having established a resident health record at the community health service center;(ii) Meeting the diagnostic and classification criteria for physical disabilities in the national diagnostic standards of Classification and Grading of Disabilities for Persons with Disabilities;(iii) Having clear consciousness, literacy skills, and the ability to communicate normally; (iv)Giving informed consent and voluntarily participating in this study.

Exclusion criteria:(i) Suffering from severe hearing or visual impairment; (ii)Being in the acute phase of an existing disease or condition;(iii) Having participated in a similar study within the past month.

### 2.2 Data Collection

All researchers underwent unified training prior to the study to ensure their sufficient understanding of the survey process. First, the researchers briefly explained the research objectives, procedures, and significance to the participants in a standardized manner. After obtaining informed consent, questionnaires were distributed on-site within the community, and participants were asked to fill them out independently in an anonymous way. If participants had difficulty understanding the questions or had visual impairments, our well - trained researchers were responsible for providing unified explanations of these items without offering any guidance or hints that could influence their self-judgment. A total of 300 questionnaires were distributed. After excluding 20 invalid questionnaires, 280 valid questionnaires were retrieved, yielding a valid response rate of 93.33%.

### 2.3 Assessment Tools

#### 2.3.1 Demographic Data Questionnaire

A self-designed questionnaire is used, which includes items such as gender, age, ethnicity, place of residence, living arrangement, educational attainment, marital status, monthly family income, personality type, medical insurance mode, types of diseases, disability level, duration of disability, self-care ability in daily life, and types of activities participated in.

#### 2.3.2 Self-rating Depression Scale

The Self-Depression Scale (SDS) was first developed by Zung in 1965 and was later revised by Wang Chunfang and others. It consists of 20 items, including 8 items for physical disorders, 8 items for depressive mental disorders, 2 items for mental and emotional symptoms, and 2 items for mental and motor disorders. The critical value of the standard score of SDS is 53 points. Scores ranging from 53 to 62 indicate mild depression, and scores ranging from 63 to 72 indicate moderate depression^[12,13]^. In this study, the Cronbach’s alpha coefficient of this scale was 0.825, indicating good internal consistency.

#### 2.3.3 Explanatory Model Interview Catalog (EMIC) Stigma Scale

The Chinese-translated version of the Explanatory Model Interview Catalog (EMIC) Stigma Scale by Chung^[9]^is a one-dimensional scale for assessing perceived stigma. It consists of a total of 15 items, and each question is measured using four options. Except for Question 2, the scores for all questions are assigned as follows: 3 points for “Yes”, 2 points for “Maybe”, 1 point for “Uncertain”, and 0 points for “No”. The scores of the 15 questions are then summed up. A higher total score indicates a stronger sense of stigma among the respondents. In this study, the Cronbach’s α coefficient of this scale is 0.831, indicating good internal consistency.

#### 2.3.4 Self-Esteem Scale

The Self-Esteem Scale was developed by Rosenberg in 1965 and later revised by Chinese scholar Wang Mengcheng^[14]^. It consists of 10 items, among which Items 3, 5, 8, 9, and 10 are reverse-scored. Tian Lumei^[15]^suggested that, considering the expression of Item 8 in the original questionnaire was inconsistent with Chinese culture, it could be forward - scored or deleted. The scale adopts the Likert four-point rating method: 1 point for “strongly disagree”, 2 points for “disagree”, 3 points for “agree”, and 4 points for “strongly agree”. The total score ranges from 10 to 40 points, and a higher score indicates a higher level of self-esteem. In this study, the Cronbach’s α coefficient of the scale is 0.770, indicating good internal consistency.

#### 2.3.5 Participation Scale

Chung^[9]^et al. translated the Chinese version of the participation scale, which consists of three factors: activity participation, social participation, and work-related participation. The scoring for each question is as follows: 1 point for no problem; 2 points for minor problems; 3 points for moderate problems; 4 points for major problems. A higher total score indicates a lower level of participation. Based on the total score, five levels are classified: 0-12 points represent no significant limitation; 13-22 points represent mild limitation; 23-32 points represent moderate limitation; 33-52 points represent severe limitation; 53-90 points represent extreme limitation. In this study, the Cronbach’s α coefficient is 0.876, indicating good internal consistency.

### 2.4 Statistical Analysis

Descriptive statistical analysis was conducted using SPSS 26.0 software. All the data in this study conformed to the normal distribution and were presented as *x*^-^ ± s. Pearson correlation analysis was employed to explore the correlations among variables. The Amos software was used to construct a structural equation model and conduct a mediation effect analysis. The model fit was evaluated according to various criteria: χ2/df<3, IFI> 0.90, CFI>0.90, TLI>0.90, SRMR<0.08, and RMSEA<0.08. If the 95% bias-corrected confidence interval (CI) based on 5000 bootstrap samples did not contain zero and *P*<0.05, the total effect, direct effect, and indirect effect were considered statistically significant.

## 3 Results

### 3.1 Common Method Bias Test

A Harman single-factor test was conducted to assess the common method bias resulting from the self-report questionnaire. The results indicated that the explanatory rate of the first principal component was 25.06%, significantly lower than the critical value standard of 40%. This suggests that there is no common method bias, and subsequent data analysis can be performed.

### 3.2 Scores of depression, self-stigma, self-esteem, and social participation among physically disabled individuals in the community

### 3.3 Differences in Depression among Community-Dwelling Persons with Physical Disabilities across Demographic Characteristics

In the demographic data, there were statistically significant differences in depression among community-dwelling persons with physical disabilities in terms of ethnicity, place of residence, educational level, monthly family income, personality type, medical insurance method, disability level, and self-care ability (*P*<0.05), as shown in Table 2.

**Table 1.** Scores of depression, self-stigma, self-esteem and social participation of community physically disabled individuals, as well as the scores of each dimension.

| Variables | Range of values (points) | $\bar{x} \pm s$ |
| --- | --- | --- |
| Depression | 0~60 | 40.94±8.27 |
| Somatic disorder and retardation | 0~18 | 12.46±2.65 |
| Depressive mood | 0~24 | 16.54±3.66 |
| Positive emotion | 0~12 | 8.25±1.89 |
| Interpersonal relationship | 2~8 | 3.69±1.33 |
| Self-stigma | 0~45 | 25.71±7.95 |
| Participation | 18~72 | 39.41±11.76 |
| Work - related participation | 3~15 | 9.00±3.16 |
| Activity participation | 7~35 | 14.60±5.17 |
| Social participation | 8~40 | 16.26±5.26 |
| Self-esteem | 10~40 | 22.44±5.14 |

**Table 2.** Comparison of Depression Scores among Community-based Physically Disabled Individuals with Different Characteristics(*n*=280,x^-^±*S*.

| Variable | Group | Score | $t/F$ | $P$ |
| --- | --- | --- | --- | --- |
| Ethnicity | Han nationality | 41.26±9.39 | 0.031 | 0.001 |
| | Other ethnicities | 45.68±8.93 | | |
| Residence | Rural area | 46.30±9.22 | 0.202 | 0.002 |
| | Urban area | 41.85±9.33 | | |
| Educational level | Primary school or below | 51.63±8.60 | 18.785 | <0.001 |
| | Junior high school | 45.01±7.82 | | |
| | High school or vocational school | 41.57±8.83 | | |
| | College or bachelor's degree | 36.37±8.55 | | |
| Monthly family income | < ¥1000 | 45.46±8.38 | 6.029 | 0.003 |
| | ¥1000~¥3000 | 41.50±8.86 | | |
| | ≥ ¥3000 | 40.98±11.74 | | |
| Personality type | Introverted | 42.31±8.62 | 7.178 | 0.001 |
| | Extroverted | 38.72±8.21 | | |
| | Neutral | 42.63±7.02 | | |
| Medical insurance type | Residents' medical insurance | 43.26±9.91 | 5.182 | 0.006 |
| | Urban employee medical insurance | 41.21±8.68 | | |
| | Self-payment | 48.91±7.93 | | |
| Disability grade | Grade 1 | 48.88±10.28 | 20.610 | <0.001 |
| | Grade 2 | 44.14±6.47 | | |
| | Grade 3 | 39.78±7.13 | | |
| | Grade 4 | 36.58±8.49 | | |
| Self-care ability | Complete self-care | 39.63±8.40 | 5.683 | 0.004 |
| | Partially self-care | 41.74±7.88 | | |
| | Unable to self-care | 46.50±7.66 | | |

### 3.4 Correlation analysis of depression, self - stigma, self-esteem, and social participation among community-dwelling physical disabled individuals

Among community-dwelling physical disabled individuals, self-stigma is significantly and positively correlated with depression (*r*=0.741, *P*<0.01) and social participation (*r*=0.728, *P*<0.01), and significantly and negatively correlated with self-esteem (*r*=-0.601, *P*<0.01).

Self-esteem is significantly and negatively correlated with both social participation (*r*=-0.613, *P*<0.01) and depression (*r*=-0.687, *P*<0.01). Social participation is significantly and positively correlated with depression (*r*=0.778, *P*<0.01). See Table 3.

**Table 3.** Correlation Analysis of Self-Stigma, Social Participation, Self-Esteem and Depression among Community-Living People with Limb Disabilities (*n*=280, *r* value)

|  | A | A1 | A2 | A3 | A4 | B | C | C1 | C2 | C3 | D |
| --- | --- | --- | --- | --- | --- | --- | --- | --- | --- | --- | --- |
| A | 1 |  |  |  |  |  |  |  |  |  |  |
| A1 | .927** | 1 |  |  |  |  |  |  |  |  |  |
| A2 | .850** | .734** | 1 |  |  |  |  |  |  |  |  |
| A3 | .883** | .715** | .674** | 1 |  |  |  |  |  |  |  |
| A4 | .687** | .529** | .487** | .560** | 1 |  |  |  |  |  |  |
| B | .703** | .661** | .579** | .640** | .444** | 1 |  |  |  |  |  |
| C | .658** | .629** | .577 ** | .582** | .370 ** | .716** | 1 |  |  |  |  |
| C1 | .552** | .531** | .490** | .485** | .298 ** | .635** | .906** | 1 |  |  |  |
| C2 | .604** | .563** | .523 ** | .541** | .375** | .667** | .894** | .720** | 1 |  |  |
| C3 | .482** | .479** | .424** | .421** | .231** | .449** | .658** | .446** | .400** | 1 |  |
| D | -.659** | -.630** | -.577** | -.578** | -.381** | -.694** | -.640** | -.566** | -.564** | -.456** | 1 |

Note: \*\* $P < 0.01$
A Depression, A1 Depressive Mood, A2 Positive Mood; A3 Physical Symptoms and Activity Lethargy; A4 Interpersonal Relationships
B Self-Stigma;
C Participation, C1 Activity Participation, C2 Social Participation, C3 Work-related Participation;
D Self-esteem.

### 3.5 Multivariate Linear Regression Analysis of Depression among Community Physically Disabled Individuals

Taking depression as the dependent variable, and the demographic data, self-stigma, self-esteem, and social participation that are significant in the univariate difference analysis as the independent variables, a multivariate linear regression analysis was conducted. The results are presented in Table 4.

**Table 4.** Multiple linear regression analysis of depression among physically disabled people in the community.

| | *B* | *SE* | $\beta$ | *t* | *p* |
| --- | --- | --- | --- | --- | --- |
| Variable | 28.72 | 3.64 | - | 7.90 | <0.001 |
| Disability grade | 1.43 | 0.40 | 0.14 | 3.60 | <0.001 |
| Self-stigma | 0.31 | 0.06 | 0.27 | 5.05 | <0.001 |
| Self-esteem | -0.52 | 0.09 | -0.28 | -6.09 | <0.001 |
| Social participation | 0.32 | 0.05 | 0.40 | 7.27 | <0.001 |

Note: $R^2=0.744$ , adjusted $R^2=0.727$ , $F=43.686$ , $P<0.05$ , “-” indicates no such data.

### 3.6 Mediating Effect Analysis and Test

AMOS 28.0 was employed to construct a Structural Equation Model (SEM) to test the hypothesized structural relationships among self-stigma, self-esteem, social participation, and depression. Prior to the mediating analysis, the goodness-of-fit indices were evaluated. The results were as follows: χ2/df=1.668, RMSEA=0.049, GFI=0.970, CFI=0.990, AGFI=0.941, NFI=0.975, and IFI=0.990. The model fit met the criteria. The results of the path analysis indicated that self-stigma had a positive predictive effect on depression (*β*=0.297, *P*<0.001); self-esteem had a negative predictive effect on depression (*β*= −0.262, *P*<0.001); social participation had a positive predictive effect on depression (*β*=0.340, *P*<0.001); self-stigma had a negative predictive effect on self-esteem (*β*= −0.404, *P*<0.001); self-stigma had a positive predictive effect on social participation (*β*=0.781, *P*<0.001); and self-esteem had a negative predictive effect on social participation (*β*=0.273, *P*<0.001). See Figure 1 for details. The Bootstrap method was used to further test the mediating effect. The original data were randomly sampled 5000 times, and the 95% confidence intervals were calculated respectively. The detailed results are presented in Table 5.

**Figure 1.**
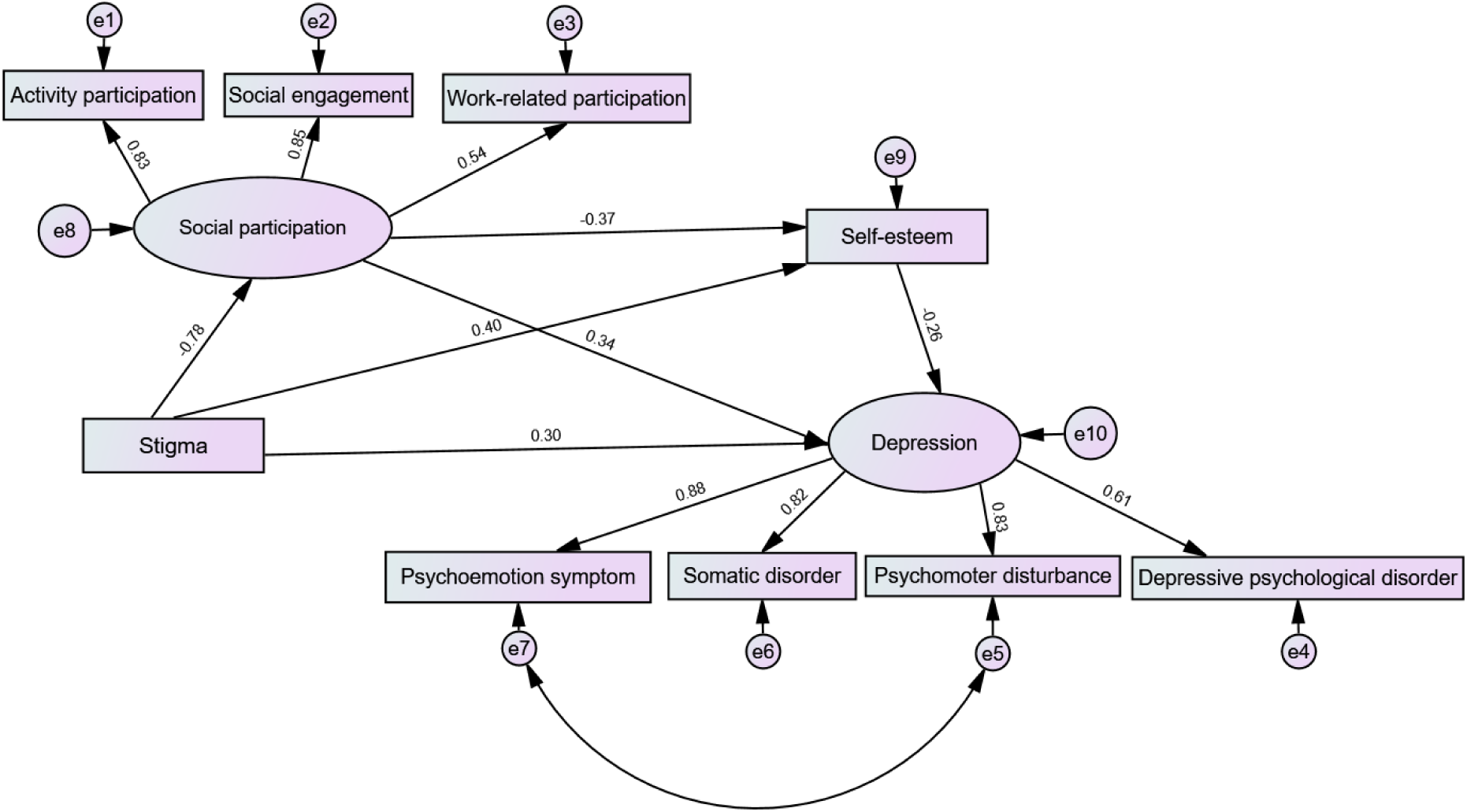
A Chain Mediating Model of the Impact of Self-stigma on Depression among Community-based Physically Disabled Individuals

**Table 5.** Results of Bootstrap Mediation Effect Test.

| Project | Estimate | SE | Z-value | Bias-Corrected |  | Percentile |  | Effect size<br>(%) |
| --- | --- | --- | --- | --- | --- | --- | --- | --- |
|  |  |  |  | 95%CI |  | 95%CI |  |  |
|  |  |  |  | Lower | Upper | Lower | Upper |  |
| Indirect<br>Effect1 | 0.265 | 0.094 | 2.820 | 0.106 | 0.482 | 0.106 | 0.480 | 35.62% |
| Indirect Effect<br>2 | 0.106 | 0.035 | 3.029 | 0.044 | 0.186 | 0.039 | 0.178 | 14.25% |
| Indirect<br>Effect3 | 0.076 | 0.028 | 2.714 | 0.033 | 0.147 | 0.030 | 0.139 | 10.22% |
| Total indirect<br>effect | 0.447 | 0.090 | 4.967 | 0.291 | 0.639 | 0.289 | 0.638 | 60.10% |
| Direct effect | 0.297 | 0.099 | 3.000 | 0.087 | 0.476 | 0.083 | 0.474 | 39.92% |
| Total effect | 0.744 | 0.034 | 21.882 | 0.671 | 0.806 | 0.671 | 0.806 | — |

Note: Indirect Effect 1: Self-stigma → Social participation → Depression Indirect Effect 2: Self-stigma → Self-esteem → Depression Indirect Effect 3: Self-stigma → Social participation → Self-esteem → Depression

## 4 Discussion

### 4.1 Analysis of the Current Situation and Influencing Factors of Depression among Community Physical Disabled Persons

The results of this study show that the depression score of physical disabled persons is (40.94±8.27), which is at an above-medium level, indicating that their mental health status is not optimistic. Introverted personality, inability to take care of oneself in daily life, low monthly family income, and high disability level are risk factors leading to depressive emotions among physical disabled persons, which is similar to previous studies^[16,17]^.

Introverted physical disabled persons are unable to actively communicate with others to regulate their emotions or express their needs when facing stressful events. Therefore, they are more likely to develop depressive emotions compared with extroverted physical disabled persons. Compared with physical disabled persons living in rural areas, urban physical disabled persons with better family economic conditions are more likely to access high-quality medical resources and health education. Ensuring personal health is beneficial for reducing the incidence of depression^[18]^. In addition, as a cultural resource, education level can increase individuals’ employment opportunities, improve their living standards, promote stronger self-control, and build a higher level of psychological security, thereby maintaining physical and mental health^[19]^.

### 4.2 The Relationship between Self-stigma and Depression

The findings of this study confirm the hypothesis that self-stigma can positively predict depression. The self-stigma scores are at a moderately high level, which is similar to the research results of Montesano^[20]^. Due to physical disabilities, they have suffered strange looks or differential treatment from others in social life. They perceive that the outside world labels them as “outsiders” and holds a stereotypical view of them as a vulnerable group. They always recognize themselves through the eyes and evaluations of others, measure their souls with a distorted yardstick, resulting in self-worth depreciation and incorrect self-perception, which in turn leads to the generation of shame^[21]^.Self-stigma can seriously damage the social functions of people with physical disabilities. When they experience a sense of stigma, they will hold a negative attitude towards themselves and engage in behaviors deviating from the social group. They may even feel disgusted with themselves, complain about the unfairness of the world, and hate their disabled state^[22]^. Subsequently, they will isolate and seclude themselves, avoid social interactions, be reluctant to have close contact with others, care more about the views and evaluations of the outside world, fear that others will know and discuss their physical disabilities, and experience increased psychological pressure, thus causing a high level of loneliness and depressive emotions^[22,23]^.

### 4.3. The Mediating Role of Social Participation between Self-Stigma and Depression

The results of this study indicate that social participation plays a mediating role between self-stigma and depression. Social participation serves as a bridge connecting individuals with society and is an important indicator reflecting public health. Social participation can positively predict the level of depression. Proactive social participation helps individuals develop a sound personality, meet their spiritual needs, realize self-worth, and increase life satisfaction. This, in turn, enhances self-esteem. When facing setbacks, individuals can attribute them correctly, adjust their emotions in a timely manner, reduce stigma internalization and social alienation, and thereby decrease the occurrence of depressive emotions^[24]^. Therefore, community workers should provide platforms and opportunities for social participation to people with physical disabilities, encourage them to actively integrate into the big social family, help them find their life goals, make them feel needed and capable of creation, and realize that they are useful to society. By doing so, it can enhance their sense of value and responsibility and reduce the generation of negative emotions.

### 4.4 The Mediating Role of Self-Esteem between Self-Stigma and Depression

This study has confirmed that self-esteem plays a mediating role between self-stigma and depression. In terror management theory, self-esteem is considered to alleviate depression.

The level of self-esteem affects an individual’s mental health, interpersonal communication, personality integrity and stability, as well as subjective initiative. High self-esteem is associated with healthy and socially acceptable behaviors, while low self-esteem contradicts socially expected cognitions and behaviors^[25]^. A high level of self-evaluation forms a positive self-attitude. When facing external negative evaluations or stressful events, individuals can respond actively to maintain their self-worth, thereby reducing the occurrence of depressive emotions. Conversely, individuals with low self-esteem are more sensitive to external remarks and are prone to depressive emotions when dealing with stressful events^[26]^. Therefore, community workers should strengthen the management of people with physical disabilities, help them build a cognitive foundation for self-esteem, guide the physically disabled group to face setbacks actively, resist psychological dilemmas, reduce the generation of negative emotions, and maintain physical and mental health.

### 4.5 The Chain Mediating Role of Social Participation and Self-Esteem between Self-Stigma and Depression

This study reveals the internal mechanism of action between self-stigma and depression among community-based individuals with physical disabilities, and further validates the chain mediating path composed of social participation and self-esteem. While expanding and modifying the application scope of the labeling theory, it also provides a more hierarchical social perspective for understanding the mental health issues of this special group. In addition, it is found that social participation is significantly correlated with self-esteem, which indicates that individuals with physical disabilities can break the double isolation at the physical and psychological levels by increasing their social participation, gradually reduce the sense of inferiority caused by stigma perception, and effectively prevent the generation and spread of negative emotions at the root. Previous studies have shown that social participation can expand the social network and social resources among people, meet individuals’ psychological dependence needs, and is more in line with the social contagion theory, that is, by enhancing social integration, individuals’ physical and mental development can be satisfied, thereby enhancing social adaptability, stimulating the social responsibility of individuals with physical disabilities, improving self-esteem levels, reducing stigma internalization and negative emotions, and being conducive to the development of mental health and the improvement of quality of life^[27,28]^. A further implication is to change the previous single assistance model for individuals with physical disabilities, construct a specific intervention system focusing on the social network and precise management of mental health of individuals with physical disabilities, empower this group, and create a social environment for equal participation to enhance the subjective well-being and self-confidence in life of this group.

### 4.6 Practical Significance

Based on the revised labeling theory, this study precisely captures the unique psychological experiences of individuals with physical disabilities in social labeling interactions. It offers a novel theoretical perspective for analyzing the internal logic of the impact of self-stigma on depression. Simultaneously, it provides solid theoretical support and practical benchmarks for subsequent targeted and scientific mental health interventions.

Community workers and rehabilitation personnel should regularly assess whether this group shows signs of self-stigma. At the cognitive level, they should actively guide and assist them in establishing a correct view of disability.

Create a “de-labeling” social interaction space for them. For example, organize activities such as social gatherings, sports meets, and various interesting competitions to encourage their active participation. Help them break free from the shackles of stigmatizing labels, reconstruct self-identity through social participation, and strengthen psychological resilience by enhancing self-esteem. Ultimately, achieve a two-way promotion of individual mental health and social civilization progress, demonstrating the humanistic warmth and practical depth of the social support system.

### 4.7 Limitations

Due to the particularity of the population, the survey subjects in this study were only physical disabled individuals from an autonomous prefecture in China, which has certain regional limitations. In the future, research can be carried out in multiple regions to make the research results more representative. This study is a cross - sectional study and cannot show the dynamic changes of research variables. In the future, longitudinal studies should be conducted to track the impact of latent variables on the depression of community-dwelling physical disabled individuals at different time points.

## 5. Conclusion

This study presents a relatively in-depth exploration of the mechanism through which self-stigma affects depression among community-dwelling individuals with physical disabilities, based on the mediating model proposed by the modified labeling theory. It confirms that self-esteem and social participation play a chained mediating role between self-stigma and depression. Therefore, community workers should not only provide routine care for the physical disabilities of these individuals but also pay attention to their psychological well-being. It is also necessary to call on the public to respect their equal right to development, promote and build a “diverse, inclusive, equal, and friendly” social ecosystem. Through the coordinated efforts of external support and internal empowerment, the goal of promoting the physical and mental health of this group can be achieved.

## CRediT Authorship contribution statement

**Hu Qianqian**: Conceptualization, Data curation, Formal analysis, Methodology, Visualization, Writing - Original Draft, Writing review & editing, Methodology. **Gong Jingjing**: Conceptualization, Data curation, Formal analysis, Supervision, Methodology, Writing - Original Draft. **Li Aixiang**: Conceptualization, Formal analysis, Visualization, Validation, Supervision. **Wang Rui**: Writing– review & editing, Visualization, Validation, Supervision, Methodology. **Chen Hui**: Conceptualization, Writing– original draft, Visualization, Formal analysis.

## Ethics approval

This study was approved by the Ethics Committee of Yanbian University. Prior to the study, all participants signed written informed consent forms after fully comprehending the study’s objectives, the voluntary nature of their participation, and the confidentiality of their responses.

## Data Availability

All relevant data are within the manuscript and its Supporting Information files.

